# Association between child sexual abuse and mid-life employment earnings

**DOI:** 10.1101/2022.01.06.22268834

**Authors:** Samantha Bouchard, Rachel Langevin, Francis Vergunst, Melissa Commisso, Pascale Domond, Martine Hébert, Isabelle Ouellet-Morin, Frank Vitaro, Richard Tremblay, Sylvana M. Côté, Massimiliano Orri, Marie-Claude Geoffroy

## Abstract

**Importance:** Individuals who have been sexually abused are at a greater risk for poor health, but associations with economic outcomes in mid-life have been overlooked.

**Objectives:** We investigated associations between child sexual abuse (≤18 years) and economic outcomes at 33-37 years, while considering type of report (official/retrospective) and characteristics of abuse (type, severity, and chronicity).

**Design:** This cohort study used data collected for the Quebec Longitudinal Study of Kindergarten Children.

**Setting:** The Quebec Longitudinal Study of Kindergarten Children is a population-based sample.

**Participants:** Participants were 3,020 boys and girls attending kindergarten in the Canadian Province of Quebec in 1986/88 and followed up until 2017.

**Main outcome/Measures:** Child sexual abuse (0-18 years old) was assessed using both retrospective self-report questionnaires and objective reports (notification to Child Protection Services). Information on employment earnings was obtained from government tax return records. Tobit regressions were used to test associations of sexual abuse with earnings adjusting for sex and family socioeconomic background.

**Results:** Of the 3,020 participants 1,320 [43.7%] self-reported no sexual abuse, 1,340 [44.3%] had no official report but were missing on the retrospective questionnaire, 340 [11.3%] reported retrospective sexual abuse, and 20 [0.7%] had official report. In the fully adjusted model, individuals who retrospectively reported being sexually abused earned US$4,031 (CI=-7,134 to -931) less per year at age 33-37 years, while those with official reports earned US$16,042 (CI=-27,465 to -4,618) less, compared to participants who were not abused. Among individuals with retrospectively reported abuse, those who experienced intra-familial abuse earned US$4,696 (CI=-9,316 to -75) less than individuals who experienced extra-familial abuse, while participants who experienced penetration earned US$6,188 (CI=-12,248 to -129) less than those who experienced non-contact abuse.

**Conclusion and Relevance:** Child sexual abuse puts individuals at risk for lasting reductions in employment earnings in adulthood. Early identification and support for sexual abuse victims could help reduce the economic gap and improve long-term outcomes.

**Key Points:** *Question:* Is child sexual abuse associated with lower mid-life employment earnings?

*Findings:* In a large population-based cohort (n=3,020), children exposed to sexual abuse had lower annual employment earnings from age 33-37 years than children nonexposed, after adjustment for childhood socioeconomic circumstances. These differences were more pronounced for individuals with official Child Protection Service reports compared to those with retrospective reports, and for individuals who experienced more severe forms of sexual abuse (i.e., penetration, intra-familial).

*Meaning:* Children exposed to sexual abuse are at risk of poor socioeconomic outcomes in mid-adulthood; interventions and support to improve long-term economic participation should be considered.

## Introduction

Child sexual abuse is a worldwide concern. Its prevalence ranges between 8 to 31% for women and 3 to 17% for men,^1^ but only a minority of sexually abused children come to the attention of Child Protection Services (CPS).^2^ Child sexual abuse can have devasting consequences on mental (e.g., depression, suicide attempts) and physical health (e.g., obesity, cardiovascular diseases) which may persist over the life course,^2-6^ and even the next generations.^7,8^ Though, less is known on the consequences of child sexual abuse on personal earnings across early adulthood and into middle-age. Clarifying the extent of these costs for individuals and society could strengthen the case for early intervention programs aimed at supporting long-term economic participation among sexual abuse victims.

To date, much of the evidence on the putative risk of sexual abuse on socioeconomic outcomes comes from studies of young adult populations. These studies reported that individuals who have been sexually abused as children had lower educational achievement,^9,10^ were more likely to be unemployed,^11^ and to benefit from welfare support in early adulthood,^12^ (18-25 years) even after accounting for childhood socioeconomic circumstances (e.g., parental earnings). Such findings raise concerns that child sexual abuse can negatively influence socioeconomic status later in life. However, to the best of our knowledge only four longitudinal studies have investigated associations between child sexual abuse and socioeconomic indicators by mid-adulthood. Three of these studies relied on self-report measures of abuse,^10,13,14^ and only one examined sexual abuse via official records, finding that those with official records of sexual abuse reported earning around US$11,000 less per year.^15^ Additionally, the studies also relied on self-report measures of socioeconomic conditions that are likely to be biased (e.g., higher attrition rates among participants with lower socioeconomic status, misreporting).^16^ Moreover, child sexual abuse may vary in severity, and it is unknown if more severe abuse puts individuals at-risk for poor economic outcomes. To our knowledge, there are no studies examining the risks of child sexual abuse using both retrospective and official reports on personal earnings using official government tax records.

### Aims

Drawing on a population-based cohort, this study examined the association between child sexual abuse and participants’ employment earnings at 33-37 years old obtained from tax return records. We investigated association for both sexual abuse officially reported to CPS (0-18 years old), and sexual abuse self-reported retrospectively at age 22 years. Additionally, given that individuals who have reported more severe forms of sexual abuse (e.g., involving penetration) might experience worse outcomes in adulthood,^17^ we investigated whether the characteristics of the sexual abuse in terms of type (i.e., intra vs. extra-familial), severity (i.e., involving penetration vs. touching/non-contact) and chronicity (i.e., one episode vs. more than one abusive episode) were associated with employment earnings among individuals with self-reported abuse. Furthermore, we estimated the risk of child sexual abuse on lost employment earnings over a 40-year working career. It is crucial to document the breadth of the consequences of child sexual abuse including on socioeconomic outcomes, especially given that poor socioeconomic status is a well-known risk factor for health and intergenerational continuity of sexual abuse.^7^

## Methods

### Participants

The Quebec Longitudinal Study of Kindergarten Children (QLSKC) is a longitudinal cohort of 3,020 boys and girls recruited while attending kindergarten in French-speaking schools in the Province of Quebec during the 1986-87 and 1987-88 school years and followed-up until 2017.^18^ Of these 3,020 children, 2,000 children (1,001 boys and 999 girls) were representative of Quebec kindergarteners (random sample) and 1,020 children (600 boys and 420 girls) were oversampled for disruptive behaviors, defined as scores ≥80th percentile on the disruptive behaviors scale from the Social Behavior Questionnaire.^19^ Ethics approval was obtained from the University of Montreal Ethics Board and Statistics Canada. Written informed consent was obtained at each assessment.

### Official report of child sexual abuse

When participants reached 18 years of age, their official reports of child sexual abuse were extracted from the Quebec Ministry of Health and CPS via the Director of Youth Protection. For the purposes of this study, we included all notifications to CPS for sexual abuse regardless of substantiation (*n*=20). Notifications for sexual abuse include (1) a situation in which the child is subjected to gestures of a sexual nature by the child’s parents or another person, with or without physical contact, and the child’s parents fail to take the necessary steps to put an end to the situation and (2) a situation in which the child runs a serious risk of being subjected to gestures of a sexual nature by the child’s parents or another person, with or without physical contact, and the child’s parents fail to take the necessary steps to put an end to the situation.^20^

### Retrospective report of child sexual abuse

At 22 years old, participants retrospectively completed five questions adapted from the *Adverse Childhood Experiences Questionnaire*^21,22^ and from the *Sexually Victimized Children Questionnaire*^23^ assessing sexual abuse before the age of 18 years. They were asked whether they had experienced unwanted sexual acts in the form of exhibitionism, sexual fondling/touching, penetration or attempted penetration by bribe, threat or force, or by drugs and/or alcohol (e.g., “Did someone show you their sexual parts or force you to show them your sexual parts when did you did not want too?”). When participants responded “yes” to any of the questions, they were asked about the nature of their relationship to the perpetrator (i.e., within the family, within the extended family, acquaintance from school, or stranger), and frequency of the abuse (e.g., one time or more). We created three variables indicating the characteristics of sexual abuse according to the (i) type of abuse: intra-familial (e.g., parents, siblings, relative) vs extra-familial perpetrator (e.g., stranger, acquaintance from school), (ii) severity: penetration/attempted penetration, sexual fondling/touching, non-contact (e.g., exhibitionism, voyeurism), and (iii) chronicity: one abusive episode vs. more than one abusive episode.

### Adult employment earnings

Employment earnings were obtained annually from 18-37 years old (1998-2017) from federal government tax returns (Statistics Canada) and linked to the QLSKC cohort.^24^ It includes pretax wages, salaries, and commissions, excluding income from capital gains. To account for random yearly variations, the mean of the 5 most recent tax return records were used (when participants were 33-37 years of age). For ease of comparability, all financial data was converted to US dollars prior to analysis using the current purchase power parity rate (CAD1=USD0.83).

### Childhood socioeconomic characteristics

Childhood socioeconomic characteristics known for their associations with child sexual abuse^12,25,26^ later socioeconomic outcomes^27^ were controlled for in all analyses. Namely, parental earnings, years of education, age at child’s birth (average of mother and father) and family composition (single vs. two parents), in addition to child’s sex and disruptive sample membership. Information on parental earnings were obtained when children were 2-7 years old (1982-1987) from government tax return records, while other characteristics were collected via a questionnaire administered to the mother when participants were aged 6 years.

### Statistical Analyses

We investigated the prevalence of sexual abuse and childhood socioeconomic correlates of child sexual abuse in the whole sample (*n*=3,020), considering both official and retrospective sexual abuse in four non-overlapping categories: (1) no official and retrospective reports; (2) no official report and missing on retrospective report; (3) retrospective report and no official report (4) official report, including seven individuals with concurrent retrospective report, and characteristics of retrospective report (type, severity, and chronicity). Next, we used tobit regressions to investigate the associations between child sexual abuse and average employment earnings in the last 5 years (33-37 years) left censured at $0. To reduce the effect of extreme outliers, income scores at or above the 99^th^ percentile were winsorized. To examine the extent to which participant sex, disruptive sample membership, and childhood socioeconomic characteristics explain the association between child sexual abuse and adulthood earnings, we fitted three models with different adjustment levels: (1) unadjusted, (2) adjusted for sex, and (3) adjusted for childhood characteristics described above (disruptive sample membership was controlled for in all models).

Missing values on childhood socioeconomic characteristics range from 0% (sex) to 30% (family composition). To avoid losing participants due to missing information on confounders, we imputed missing values using multiple imputation by chained equations;^28^ analyses were conducted across 50 pooled imputed datasets.

To estimate the long-term economic consequence of child sexual abuse, we estimated the loss of individual earnings over a 40-year work career:

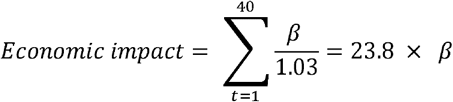

Where *B* is the estimate from the fully adjusted model, while assuming a discount rate of 3%.^29^ This indicates the lifetime loss of earnings attributable to sexual abuse, independently from background socioeconomic characteristics.

All analyses were repeated in regard to the type of abuse, severity, and chronicity of abuse as exposure among the individuals with retrospective report of abuse.

## Results

Frequencies of child sexual abuse and characteristics of abuse are presented in **Table 1**. Of the 3,020 participants 1,320 [43.7%] self-reported no sexual abuse, 1,340 [44.3%] had no official report but were missing on the retrospective questionnaire, 340 [11.3%] reported retrospective sexual abuse, and 20 [0.7%] had official report. While excluding the 1,340 participants with missing information, 25.7% of the sample reported child sexual abuse.

**Table 1.** Prevalence and characteristics of child sexual abuse

| ***Child sexual abuse, No. (%)*** | **Total**<br>***n* = 3020** |
| --- | --- |
| No sexual abuse | 1320 (43.7) |
| Missing retrospective report, but no official report | 1340 (44.3) |
| Retrospectively reported sexual abuse | 340 (11.3) |
| Officially reported sexual abuse | 20 (0.7) |
| ***Characteristics of retrospectively reported child sexual abuse, No. (%)*** | ***n* = 350** |
| ***Type of abuse*** |  |
| Intra-familial | 130 (38.6) |
| Extra-familial | 210 (61.4) |
| ***Severity of abuse*** |  |
| Non-contact | 80 (21.6) |
| Fondling | 130 (38.0) |
| Penetration or attempted penetration | 140 (40.3) |
| ***Chronicity*** |  |
| One episode | 290 (82.4) |
| More than one episode | 60 (17.6) |

Note. In accordance with Statistics Canada data protection requirements, displayed counts are rounded to base 10.

With regards to characteristics of abuse, more participants self-reported extra-familial (61.4%), penetration or attempted penetration (40.3%) and fondling (38.0%) and were abused on one occasion (82.4%). Participants who were sexually abused or exposed to the most severe forms of abuse were more likely to come from an underprivileged family in terms of parental education and earnings than those who were not sexually abused **(eTable 1 and eTable 2)**.

Descriptive statistics for the association between child sexual abuse and employment earnings from 18-37 years are reported in **Figure 1 and Figure 2**. The average of the last 5 available years (ages 33–37) was US$32,800 (*SD*=24,840). Participants who experienced sexual abuse earned less than those who did not experience sexual abuse, with official reports being associated with the lowest earnings across follow-up. Analyses controlling for sex, disruptive behaviour and childhood socioeconomic characteristics including parental earnings are shown in **Table 2**. Compared to those who did not experience child sexual abuse, individuals who retrospectively reported sexual abuse earned US$4,031 (CI =-7,134;-931) less, while those with official reports earned US$16,042 (CI =-27,465;-4,618) less. Those missing the retrospective report, but with no official report, also had lower annual earnings, but this difference was not statistically significant US$-1,906 (CI =-3,912;98). Over a 40-year working career, the loss of personal earnings attributable to sexual abuse was US$95,956 (CI =-169,740; - 22,171) for retrospectively reported abuse and US$381,800(−653,709;-109,946) for officially reported abuse. This estimates account for all confounders used in the fully adjusted model, and a 3% annual discount rate.

**Table 2.** Associations of officially and retrospectively reported child sexual abuse and adult employment earnings (33-37 years old); *n* = 3020.

|  | Differences in employment earnings<sup>a</sup> |  |  |
| --- | --- | --- | --- |
|  | Unadjusted<sup>b</sup> | Adjusted for sex<sup>b</sup> | Adjusted for family socioeconomic characteristics<sup>b,c</sup> |
|  | *B* [95%CI] | *B* [95%CI] | *B* [95%CI] |
| Missing retrospective report, but no official report | **-3,181** [-5,236 to -1,127] | **-4,132** [-6,157 to -2,107] | -1,906 [-3,912 to 98] |
| Retrospectively reported sexual abuse | **-9,107** [-12,274 to -5940] | **-5,513** [-8,688 to -2,336] | **-4,031** [-7,134 to -931] |
| Officially reported sexual abuse | **-26,993** [-38,929 to -15,055] | **-23,284** [-35,000 to -11,570] | **-16,042** [-27,465 to -4,618] |

Note.<sup>a</sup>Based on imputed values ( $N = 3020$ ).
<sup>b</sup>Estimates are beta coefficients from Tobit regression with 95% confidence intervals and indicate amounts in US dollars; all three models are adjusted for cohort membership (i.e., disruptive vs. representative sample)
<sup>c</sup>Adjusted for family characteristics including child sex, maternal and paternal education, parental earnings, parental mean age at childbirth, and family composition.
Reference category = no abuse; significant differences at $p < 0.05$ are bold.

**Figure 1.**
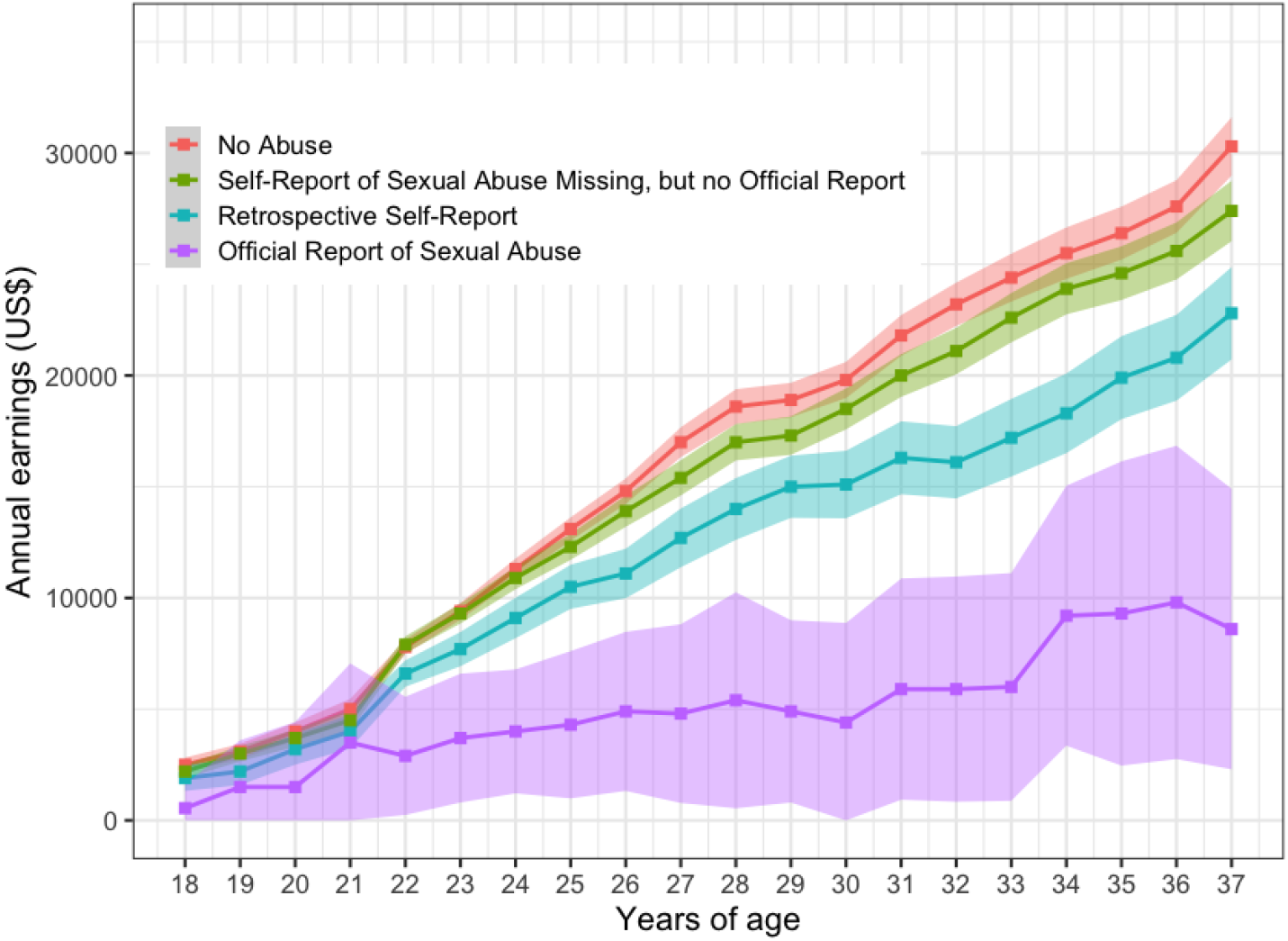
Descriptive statistics for employment earnings from 18 to 37 years old by sexual abuse group, *n* = 3,020^a^ Note. ^a^In accordance with Statistics Canada data protection requirements, displayed counts are rounded to base 10 and employment earnings were rounded to the closest 100.

**Figure 2.**
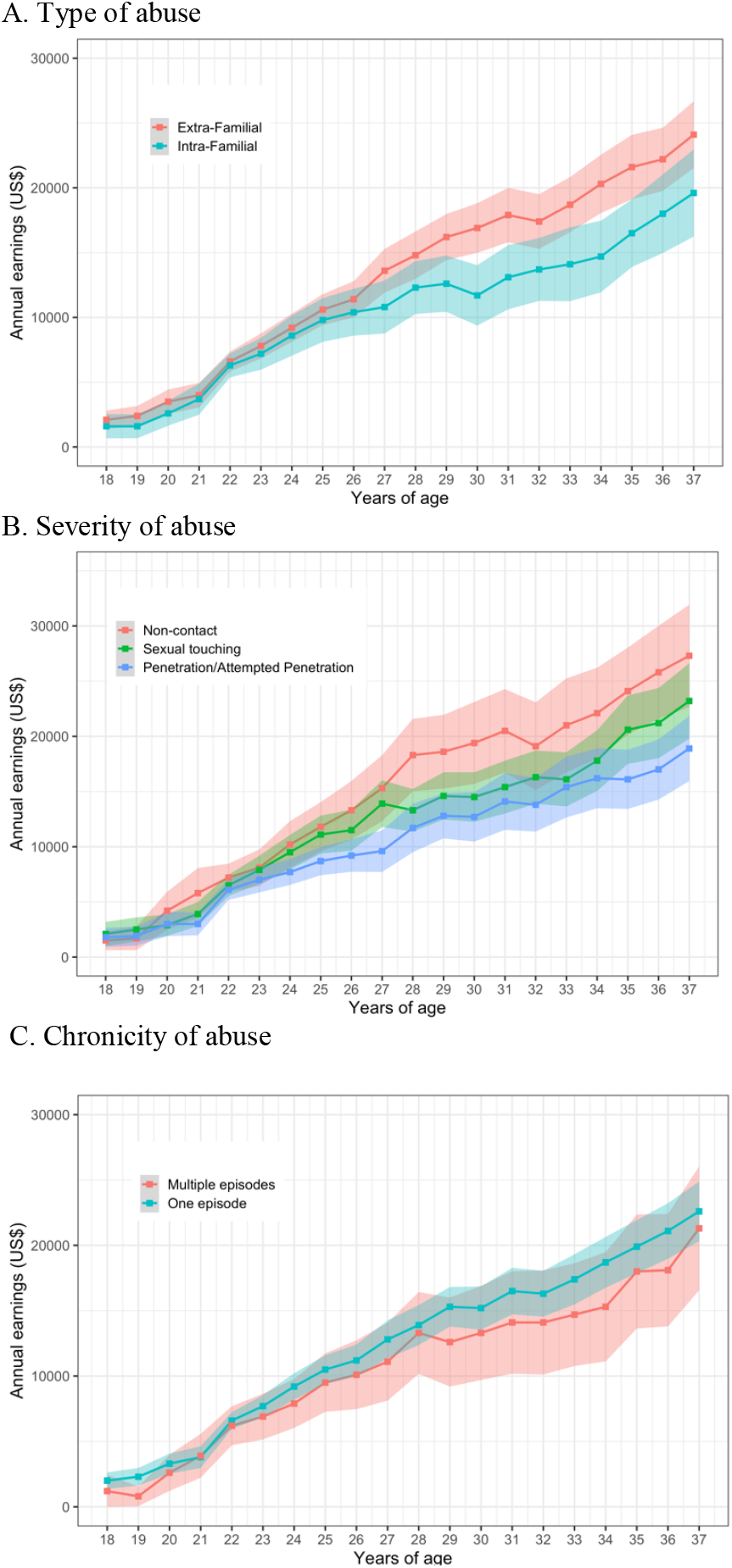
Descriptive statistics for employment earnings from 18 to 37 years (*n* = 350)^a^ Note. ^a^In accordance with Statistics Canada data protection requirements, displayed counts are rounded to base 10 and employment earnings are rounded to the closest 100.

As illustrated in **Figures 2a and 2b**, participants who experienced intra-familial abuse or penetration/attempted penetration earned less from 18-37 years old than those who experienced extra-familial abuse or non-contact sexual abuse, respectively (no difference was observed for chronicity of abuse, see **Figure 2c**). Fully adjusted analyses revealed that compared to those who experienced extra-familial abuse, individuals who experienced intra-familial abuse earned US$4,696 (CI=-9,316;-75) less. Individuals who experienced more severe sexual abuse (i.e., penetration) earned US$6,188 (CI=-12,248;-129) less than those who experienced non-contact sexual abuse. No significant difference in earnings was observed between participants who experienced sexual touching vs. non-contact sexual abuse and those with one episode vs. multiple episodes of sexual abuse (**Table 3)**. Results based on complete cases did not substantively differ from the main results (**eTable3 and eTable4)**. Across a 40-year working career, the loss of personal earnings attributed to severity of sexual abuse (e.g., penetration) was US$147,309 (CI = -291,532;-3,086). The loss of personal earnings attributed to intra-familial abuse was US$111,804 (CI = -221,741;-1867).

**Table 3.** Associations of characteristics of retrospectively reported child sexual abuse among victims of sexual abuse adult employment earnings; *n* = 350.

|  | Differences in employment earnings<sup>a</sup> |  |  |
| --- | --- | --- | --- |
|  | Unadjusted<sup>b</sup> | Adjusted for sex<sup>b</sup> | Adjusted for family socioeconomic characteristics<sup>b,c</sup> |
| ***Type of abuse*** |  |  |  |
| Intra-familial<br>(vs. extra familial abuse) | **-6,898**<br>[-11,823 to -1,973] | **-6,416**<br>[-11,275 to -1,557] | **-4,696**<br>[-9,316 to -75] |
| ***Severity of abuse*** |  |  |  |
| Sexual touching<br>(vs. non-contact) | -5,603<br>[-11,934 to 726] | -4,502<br>[-10,813 to 1807] | 3,515<br>[-9,456 to 2,427] |
| Penetration<br>(vs. non-contact) | **-10,471**<br>[-16,764 to -4,179] | **-8,517**<br>[-14,916 to -2,118] | **-6,188**<br>[-12,248 to -129] |
| ***Chronicity*** |  |  |  |
| Multiple episodes<br>(vs. one episode) | -3,010<br>[-9,414 to 3,391] | -1,518<br>[-7,886 to 4,847] | -2,081<br>[-8,056 to 3,892] |

Note. <sup>a</sup>Based on imputed values ( $N = 350$ ).
<sup>b</sup>Estimates are beta coefficients from Tobit regression with 95% confidence intervals and indicate amounts in US dollars; all three models are adjusted for cohort membership (i.e., disruptive vs. representative cohorts).
<sup>c</sup>Adjusted for family characteristics including child sex, maternal and paternal education, parental earnings, parental mean age at childbirth, and family composition.
Significant differences at $p < 0.05$ are bold.

## Discussion

To our knowledge, this is the first longitudinal study to investigate associations between child sexual abuse (retrospective and official reports) and long-term employment earnings by mid-life. Using employment earning data obtained from government tax return records linked to over 3000 participants from the community, we found that individuals who experienced child sexual abuse endured poor long-term economic outcomes. To illustrate, individuals who retrospectively reported being exposed to penetrative and intra-familial sexual abuse had over US$100,000 lower annual earnings than non-abused children across a 40-year working career, while those who were notified to CPS for sexual abuse had over US$300,000 lower annual earnings than non-abused children across their careers. Our estimates were robust to adjustment for a range of childhood confounders including parental earnings gathered from tax records.

### Methodological considerations

Our study has several strengths. First, we relied on a large population-based cohort followed for almost four decades. Second, we used both retrospective and official CPS records to assess child sexual abuse. By using official records, we obtained a more objective rating of child sexual abuse and avoid many of the reporting biases (e.g., desirability or memory biases) found in retrospective data.^30,31^ Alternatively, by using retrospective reports of child sexual abuse, it allows us to anonymously collect large amounts of sensitive data,^30^ while capturing cases that have gone unreported to CPS.^32^ Indeed, within the Province of Quebec, only 0.48 incidents for every 1,000 children living in the province were reported to CPS.^33^ Third, for those with retrospective reports, we had detailed information on the characteristics of sexual abuse allowing us to examine how type, severity and chronicity influence employment earnings. Fourth, we used objective measures of employment earnings (i.e., government tax returns). Finally, we controlled for key childhood confounders including parental earnings which were also obtained from tax records.

However, our findings also need to be interpreted in light of methodological limitations. First, as about one-third of participants did not answer the retrospective questionnaire on sexual abuse, we might have underestimated the true association between retrospectively reported sexual abuse and employment earnings. But it is worth noting that our main analyses did not reveal differences between participants who were missing on the retrospective questionnaire and those who were not abused in terms of employment earnings. Second, despite our large sample, only 20 participants were identified by CPS for sexual abuse, increasing confidence intervals around estimates, and preventing us from completing detailed analyses regarding characteristics of the abuse and the victim (e.g., sex). Therefore, a replication of our study with an at-risk sample for sexual abuse is recommended. Third, we did not control for other forms of child maltreatment (e.g., physical abuse, neglect), hence we do not know whether the earning difference is solely due to child sexual abuse or whether it could be better explained by the cumulative risk of different forms of abuse and childhood adversity. Fourth, we did not control for child (before the abuse) and parental psychopathology, which could increase sexual abuse risk and lower long-term earnings. Finally, we were unable to examine the differential risk by sex due to a lower number of victims among men.

Our results concur with some previous studies,^10,14^ but not others. For example, a study based on the Ontario Child Health Study (N=1,454) failed to report an association between child sexual abuse, assessed retrospectively with one self-reported item and self-reported earnings among employed adults at 21-35 years.^13^ In our study, although there was an overall association between retrospectively reported child sexual abuse and employment earnings, additional analyses revealed that associations with employment earnings were seen only for the most severe forms of sexual abuse, namely penetration, intra-familial abuse, and official reports, which could not be tested in the previous study using a single broad self-reported item to document abuse. To illustrate, participants abused by a family member earned over US$4,000 less between the ages of 33-37 per year than those abused by someone outside of their family. Such findings are in line with a prior study within this cohort showing that suicide attempt risk at 22 years old was 5 times higher for those sexually abused by a family member compared to those abused by someone outside the family.^34^

In our study, the largest earning differences was observed for children with official reports of sexual abuse, with mean individual annual earnings over US$16,000 less (between ages 33-37 years old) compared to their non-abused counterparts. As only 20 participants had official reports of sexual abuse, these results should be interpreted with caution. Nevertheless, another prospective cohort of 807 males and females with official records of sexual abuse and self-reported socioeconomic outcomes found that sexual abuse was associated with an income gap of US$8,000 at age 41, and that abused individuals were less likely to be in a skilled job, employed, own stocks, a vehicle or a home.^15^ Typically, children who are reported to CPS for sexual abuse represent the most severe instances of sexual abuse^35^ which often arise in combination with other forms of abuse (e.g., neglect, physical abuse) and childhood adversity.^33,36^

As most prior studies, associations between sexual abuse and personal earnings were robust to adjustment of key confounding factors, including parental household earnings. However, in an article using data from the Christchurch Health and Development Study,^12^ the association between child sexual abuse and gross income was lost after adjustment for childhood factors.

### Potential Underlying Mechanisms

Future studies are needed to replicate our findings but also to elucidate intermediary pathways. Deficits in cognitive abilities and educational attainments,^37^ poor physical (e.g., obesity,^4^ gastrointestinal symptoms,^38^ headaches^39^) and mental health (e.g., impairing depression,^6^ anxiety,^40^ substance use problems,^38^ suicide attempts^29^) associated with child abuse could be potential mediating pathways. Of note, in the 1958 British Birth Cohort associations of retrospective report of child abuse and financial insecurity and income-related support at age 50 years were not mediated by cognitive abilities and mental health during adolescence.^10^

### Implications and conclusions

Overall, our findings suggest that child sexual abuse may contribute to socioeconomic inequalities in mid-adulthood. Such inequalities were especially pronounced for the most severe forms of abuse (official reports to CPS, intra-familial abuse, and penetration). As earnings is a marker for healthy aging,^41^ longevity,^42^ and can protect against the intergenerational continuity of child sexual abuse and maltreatment^43^ helping victims of sexual abuse to maximise their economic potential would contribute to resilient functioning and yield important social and economic returns for individuals and potentially society.

## Data Availability

All data produced in the present work are contained in the manuscript

---

**eTable 1:** Childhood socioeconomic characteristics by officially and retrospectively reported child sexual abuse.

|  | No sexual abuse<br>(*n*=1320) | Missing<br>retrospective<br>report, but no<br>official report<br>(*n*=1340) | Officially reported<br>sexual abuse<br>(*n*=20) | Retrospectively<br>reported sexual<br>abuse<br>(*n*=340) | P-values for<br>group<br>differences |
| --- | --- | --- | --- | --- | --- |
|  | *M (SD)* |  |  |  |  |
| **Characteristics** |  |  |  |  |  |
| Maternal education (years) | 12.17 (2.60) | 11.21 (2.62) | 10.15 (2.08) | 11.84 (2.42) | <.001 |
| Father's education (years) | 12.29 (3.34) | 11.29 (3.38) | 9.62 (3.16) | 12.22 (3.48) | <.001 |
| Parental mean age (years) | 27.93 (4.24) | 27.69 (4.62) | 26.37 (5.13) | 27.93 (4.39) | .236 |
| Parental earnings in US dollars<sup>a</sup> | 27,100 (13970) | 22,600 (15510) | 10,800 (10190) | 23,400 (14380) | <.001 |
| Family composition (single parent); No.(%) | 119 (5.6) | 113 (5.3) | <5 | 32 (1.5) | .987 |

Note. Based on a maximum N ranging from 2136 to 2998.
<sup>a</sup>Rounded to the closest 100 according to Statistics Canada data protection regulations.

**eTable 2:** Childhood socioeconomic characteristics by officially by characteristics of retrospectively reported child sexual abuse among victims of sexual abuse.

| Characteristics | Type of abuse |  |  | Severity of abuse |  |  |  | Chronicity |  |  |
| --- | --- | --- | --- | --- | --- | --- | --- | --- | --- | --- |
|  | Intra-familial | Extra familial | P-values for group differences | Non-Contact | Sexual touching | Penetration | P-values for group differences | One episode | More than one episode | P-values for group differences |
|  | *M (SD)* |  |  |  |  |  |  |  |  |  |
| Maternal education (years) | 11.54 (2.32) | 11.93 (2.52) | .158 | 12.00 (2.46) | 11.84 (2.60) | 11.61 (2.30) | .527 | 11.78 (2.50) | 11.78 (2.23) | .995 |
| Father's education (years) | 11.48 (3.04) | 12.06 (3.70) | .007 | 12.64 (3.53) | 12.46 (3.70) | 11.67 (3.22) | .110 | 12.20 (3.52) | 12.10 (3.43) | .842 |
| Parental mean age (years) | 27.55 (4.38) | 28.13 (4.51) | .248 | 28.33 (4.04) | 27.66 (4.21) | 27.92 (4.90) | .586 | 27.84 (4.42) | 28.25 (4.67) | .510 |
| Parental earnings in US dollars<sup>a</sup> | 21,300 (13,440) | 24,300 (14,930) | .057 | 26,000 (15,810) | 24,100 (13,860) | 20,700 (14,430) | .022 | 23,000 (14,630) | 23,800 (13,510) | .670 |
| Family composition (single parent); No.(%) | 13 (39.4) | 20 (60.6) | .847 | 6 (18.2) | 11 (33.3) | 16 (48.5) | .712 | 27 (81.8) | 6 (18.2) | .709 |

Note. Based on a maximum N ranging from 257 to 350.
<sup>a</sup>Rounded to the closest 100 according to Statistics Canada data protection regulations.

**eTable 3:** Associations of child sexual abuse with adulthood employment earnings, completed case, *n* = 1740.

|  | Differences in employment earnings<sup>a</sup> |  |  |
| --- | --- | --- | --- |
|  | Unadjusted<sup>b</sup> | Adjusted for sex<sup>b</sup> | Adjusted for family socioeconomic characteristics<sup>b,c</sup> |
| ***Personal earnings*** |  |  |  |
| Missing retrospective report, but no official report | **-3,389** [-5,458 to -1,323] | **-4,382**<br>[-6,412 to -2,350] | 22.95<br>[-2,596 to 2,641] |
| Retrospectively reported sexual abuse | **-9,148**<br>[-12,339 to -5,960] | **-5,375**<br>[-8,568 to -2,181] | -3,375<br>[-7,385 to 634] |
| Officially reported sexual abuse | **-26,978**<br>[-38,970 to -14,985] | **-23,174**<br>[-34,923 to -11,426] | **-16,515**<br>[-32,827 to -203] |

<sup>a</sup>Based on non-imputed values ( $N = 1740$ ).
<sup>b</sup>Estimates are beta coefficients from Tobit regression with 95% confidence intervals and indicate amounts in US dollars; all three models are adjusted for cohort membership (i.e., disruptive vs. representative cohorts).
<sup>c</sup>Adjusted for family characteristics including child sex, maternal and paternal education, parental earnings, parental mean age at childbirth, and family composition. Reference category = no abuse; significant differences at $p < 0.05$ are bold.

**eTable 4:** Associations of characteristics of retrospectively reported child sexual abuse among victims of sexual abuse with adulthood employment earnings, completed case, *n* = 200.

|  | Differences in employment earnings in US dollars<sup>a</sup> |  |  |
| --- | --- | --- | --- |
|  | Unadjusted<sup>b</sup> | Adjusted for sex<sup>b</sup> | Adjusted for family socioeconomic characteristics<sup>b,c</sup> |
| **Type of abuse** |  |  |  |
| Intra-familial (vs. extra familial abuse) | **-6,886**<br>[-11,781 to -1,992] | **-6,297**<br>[-11,117 to -1,478] | -1,823<br>[-10,539 to 2,010] |
| **Severity of abuse** |  |  |  |
| Sexual touching (vs. non-contact) | -5,719<br>[-11,881 to 710] | -4,401<br>[-10,670 to 1,867] | **-8,479**<br>[-16,506 to -450] |
| Penetration (vs. non-contact) | **-10,783**<br>[-17,055 to -4,513] | **-8,587**<br>[-14,960 to -1,971] | **-10,357**<br>[-18,362 to -2,351] |
| **Chronicity** |  |  |  |
| Multiple episodes (vs. one episode) | -3,375<br>[-9,721 to 2,968] | -1,699<br>[-8,004 to 4,604] | -2,706<br>[-10,939 to 5,525] |

<sup>a</sup>Based on non-imputed values ( $N = 200$ ).
<sup>b</sup>Estimates are beta coefficients from Tobit regression with 95% confidence intervals and indicate amounts in US dollars; all three models are adjusted for cohort membership (i.e., disruptive vs. representative cohorts).
<sup>c</sup>Adjusted for family characteristics including child sex, maternal and paternal education, parental earnings, parental mean age at childbirth, and family composition. Significant differences at $p < 0.05$ are bold

